# Non-operative management of uncomplicated appendicitis in children: a randomised, controlled, non-inferiority study evaluating safety and efficacy

**DOI:** 10.1101/2023.07.27.23292746

**Authors:** Susan Elizabeth Adams, Meegodage Roshell Swindri Perera, Saskia Fung, Jordon Maxton, Jonathan Karpelowsky

## Abstract

**Background:** Appendicitis is the commonest paediatric surgical emergency. Adult studies suggest non-operative management (NOM) may have a place in care. There have been no adequately powered randomized controlled trials in children. Objective: to determine the safety and efficacy of NOM for paediatric simple appendicitis.

**Methods:** A non-inferiority randomized controlled trial was conducted comparing operative (OM) to NOM of SA in children aged five-15 years. Primary outcome was treatment success (no unplanned or unnecessary operation, or complication) at 30 days and 12 months, with a non-inferiority margin of 15%. (anzctr.org.au:ACTRN12616000788471).

**Results:** From 11 June 2016 to 30 November 2020, 222 children were randomized: 94 (42·34%) to OM and 128 (57·66%) to NOM. Non-inferiority of NOM was not demonstrated at either time point, with 45.67% of NOM patients subsequently undergoing operation. There was no significant difference in complications (p=0.399).

**Conclusions:** While noninferiority was not shown, NOM was safe, with no difference in adverse outcomes between the two groups. Further research to refine the place of NOM of simple appendicitis in children is required, including nuanced patient selection, longer term evaluation, the place of choice, and the acceptability of the treatment for children and their carers.

**Level of evidence:** Level 1

**Highlights:** *What is currently known:* There have been multiple studies examining the role of non-operative management (NOM) with antibiotics alone for appendicitis in adults, with inconclusive evidence regarding its non-inferiority to operative management. There have only been three pilot randomized control trial to date in children.

*What this study adds:* This is the first adequately powered randomized controlled trial in children. While non-inferiority was not shown, non-operative management was safe, and associated with significantly shorter time away from school and usual activities.

## Background

Appendicitis is one of the most common paediatric surgical emergencies, with a lifetime risk of 7-8%,^1^ a childhood risk of 2.5%,^2^ and paediatric population prevalence in New South Wales of 1:700.^3^ Its frequency, associated morbidity,^2^ measurable mortality^4^ and financial costs^3^ of surgical management, mean the condition places a considerable burden on paediatric health systems.

Timely operative management (OM) of simple appendicitis (SA) has remained the cornerstone of management, the central tenant being prevention of advancement to complicated appendicitis (CA). However, there is evidence that complicated appendicitis can be considered a separate pathological entity, not necessarily the inevitable consequence of delayed operation.^5^

In recent years, studies in adults have examined the role of non-operative management (NOM) of SA.^6-12^Rollins’ 2016^13^ and Prechal’s 2019^14^ meta-analyses of the same five prospective randomized controlled trials,^7,9-12^ reported 62.5% antibiotic treatment efficacy at one year. Prechal (2019) calculated complications to be non-significantly different, concluding appendicectomy should be first line treatment, while Rollins (2016) calculated an overall 48% risk reduction for NOM, seeing it as viable, safe, and effective.

The evidence in children is heterogenous.^15^ While there are published protocols planning for large studies,^16-18^there have only been three small randomized controlled trials,^19-21^reporting an appendicectomy rate after NOM of 7-59%. In the most recent systematic review and meta-analysis of 21 disparate publications,^22^ 92% of studies concluded that NOM was effective, with an eventual appendicectomy rate of 24% overall but up to 60% in one study.^23^

There is a need for further well designed, adequately powered, prospective clinical trials to determine the place of NOM in managing paediatric SA.

### Aims

The primary objective was to compare OM and NOM of clinically diagnosed SA in children with regards to efficacy and safety. The hypothesis was that NOM is not inferior and that it is a safe alternative to OM for SA.

## Methods

A multicentre, prospective, open label, non-inferiority randomized controlled trial (anzctr.org.au: ACTRN12616000788471) was conducted between 11 June 2016 and 30 November 2020, at Sydney Children’s Hospital and The Children’s Hospital at Westmead, Sydney, Australia. The two hospitals see approximately 92,000 emergency presentations and 600 cases of acute appendicitis annually. There were two parallel treatment groups – OM and NOM. The study design was chosen because there is no evidence that NOM is superior, and because blinding was not possible.

The SPIRIT checklist (2013) was used to draft the protocol, published in 2016.^24^ Salient points and changes are summarised here and in Figure 1. Ethical approval was obtained from the Sydney Children’s Hospitals Network Ethics committee (HREC/15/SCHN/266).

**Figure 1.**
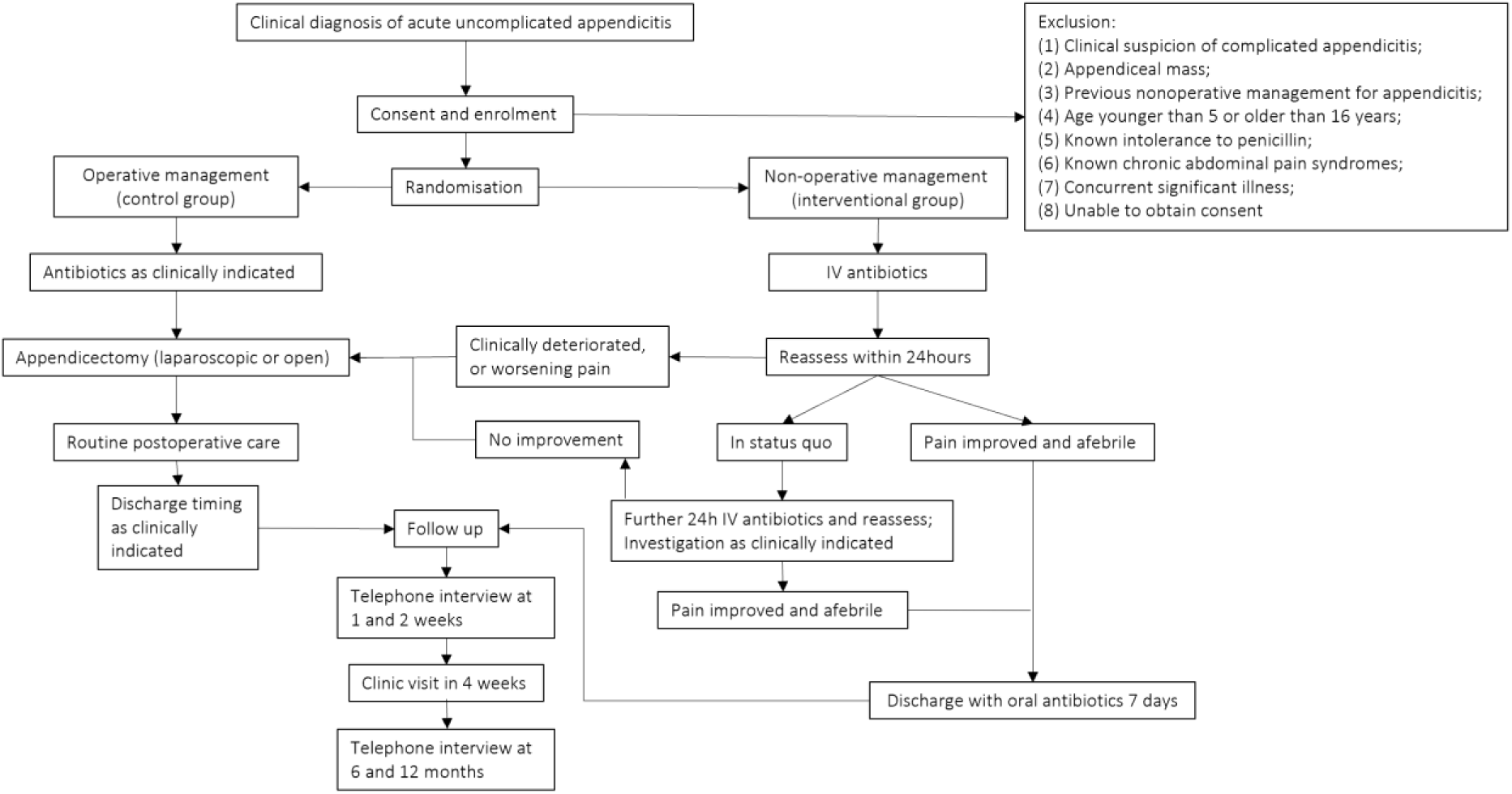
Study design.

### Eligibility

Patients aged five-15 were eligible if diagnosed with SA on clinical findings, +/- laboratory and imaging results that before the study would have led to the recommendation to operate. Exclusion criteria are summarised in Figure 1.

### Recruitment and randomization

The surgical team invited participation, with appropriate verbal and written information. Computer generated sealed envelope randomization to OM or NOM was performed at the start of the study by the senior author JK, as described in the published protocol.^24^ Randomization was repeated during the study for groups where all sealed envelopes had been used.

### Interventions

#### Antibiotics

Antibiotic use and duration in each arm is outlined in Figure 1. From 11 June 2016, the NOM group received intravenous piperacillin with tazobactam (Tazocin©), eight-hourly at 100mg/kg/dose. A world-wide shortage led to trial suspension on 17 July 2017, recommencing on 26 March 2018 once the hospitals’ antibiotic stewardship teams selected intravenous amoxicillin with clavulanate acid (Augmentin©), eighth-hourly at 25mg/kg/dose as a replacement. Children were discharged on twice daily oral Augmentin©, 22.5mg/kg/dose.

#### Operation

Children in the OM arm were managed as per usual practice. In the NOM group, appendectomy was performed if the patient’s condition worsened at any time, they were not well enough for discharge at 48 hours, or if they re-presented to hospital at any time during the study period with symptoms consistent with acute appendicitis.

#### Variables

The main explanatory variable was treatment group: OM or NOM. The primary outcome was treatment success, measured at 30 days and 12 months, based on occurrence of (a) an unplanned or unnecessary operation, and (b) complications of the appendicitis itself in the NOM arm or due to the management of the appendicitis in the OM arm. An unplanned operation was defined as an operation in a child randomized to NOM, or an additional operation in a child randomized to OM. An unnecessary operation included removal of an appendix that was not inflamed on histological examination. Complicated appendicitis was defined as appendicitis with perforation.

Safety was assessed based on complications, including: CA in the non-operative arm; bowel obstruction; surgical site infections; peritonitis; abscess or phlegmon formation; and sepsis. Other secondary outcomes are listed in Table 2.

## Statistical analysis

Sample size calculation and non-inferiority rationale are explained in the original protocol.^24^ Treatment success of 90% was expected for OM.^7,25-27^A success rate for NOM of less than 75% was considered unacceptably low, giving a non-inferiority margin of 15%. Requiring a sample size of 160, it was planned to recruit 110 patients per group to allow for up to 25% loss to follow up. Due to slow enrolment, and higher than expected participant retention, recruitment was halted when the OM arm reached 94.

Categorical variables were characterised with frequency and percentage and compared using Chi Square or Fisher’s exact test where numbers were <5. Continuous variables were described with mean (standard deviation) if normally distributed, otherwise with median (interquartile range). They were compared using independent sample t-test if normally distributed; and Mann-Whitney test if not. A p-value of <0·05 was considered significant.

Analysis was performed using IBM SPSS Statistics for Windows, version 27·0, 2020, and R version 4.2.2 (package “TOSTER”). Comparative analysis was done on an intention to treat (ITT) basis with adjustment for missing data using Holm’s step-down Bonferroni method. Non-inferiority of NOM was examined using a two one-sided test (TOST) from the TOSTER ‘TOSTtwo.prop’ function.^28^ The null-hypothesis of inferiority was **rejected** if the lower bound of the upper tail 95% one-sided confidence interval (CI) for the mean success proportion difference lay at -0.15 or higher. The function also tested the null hypothesis of non-inferiority, **rejected** if the upper bound of the lower tail 95% CI for the mean success proportion difference lay at 0.15 or lower.

## Results

During the study period, 1,570 cases of appendicitis presented, of which 222 eligible children were enrolled: 128 (57·66%) randomized to NOM and 94 (42·34%) to OM. (Figure 2) The two groups were comparable on all explanatory variables (Table 1). The median PAS was seven (IQR: 6.0-8.0), indicating a high probability of appendicitis.^29^ Non-inferiority was not demonstrated at 30 days or 12 months (Table 2, Figure 3): for both, the treatment success proportion difference and lower bounds of the upper tail 95% CI lay below the non-inferiority margin of -0.15. The upper bound of the lower tail 95% CI lay above -0.15 at 30 days but below at 12 months, meaning NOM was significantly inferior to OM by that time. Comparing the two antibiotic eras, treatment success was no different. [OM: Tazocin©: 28/32 (87.50%) vs Augmentin© 54/59 (91.52%). OR 1.543 (0.38 - 6.21), p=0.715. NOM: Tazocin© 26/52 (50.00%) vs Augmentin© 42/75 (56.00%) OR 1.273 (6.26 - 2.587), p=0.505]

**Figure 2:**
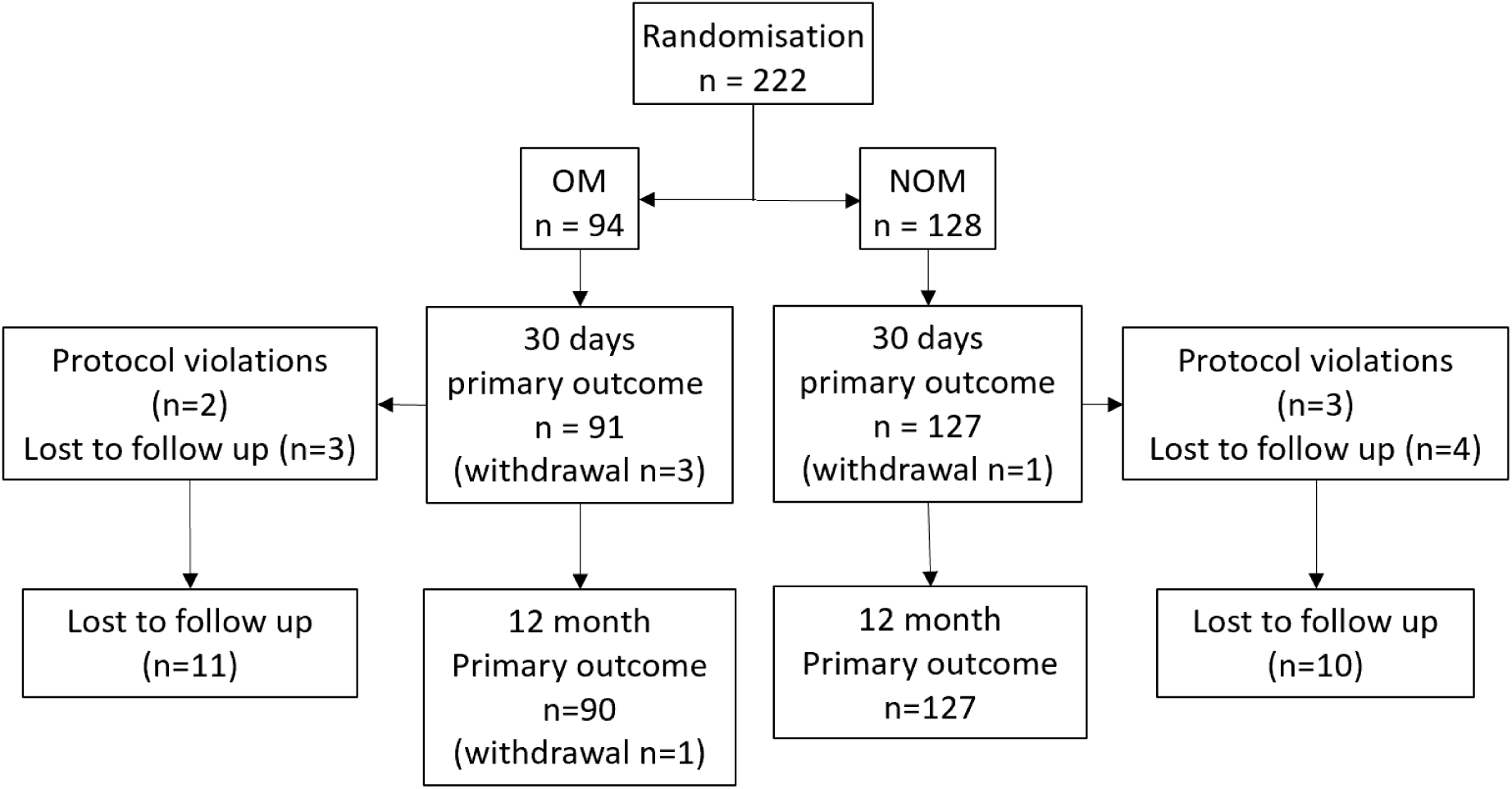
Study flow diagram. Note. Protocol violations a) operative arm: one penicillin allergy and one patient recovering on pre-operative antibiotic therapy; b) non-operative arm: two penicillin allergies, and one alternative antibiotic prior to enrolment. Lost to follow up: not contactable at clinical follow-up or by telephone. Withdrawals excluded from analysis. Lost to follow-up adjusted for in final analysis.

**Figure 3:**
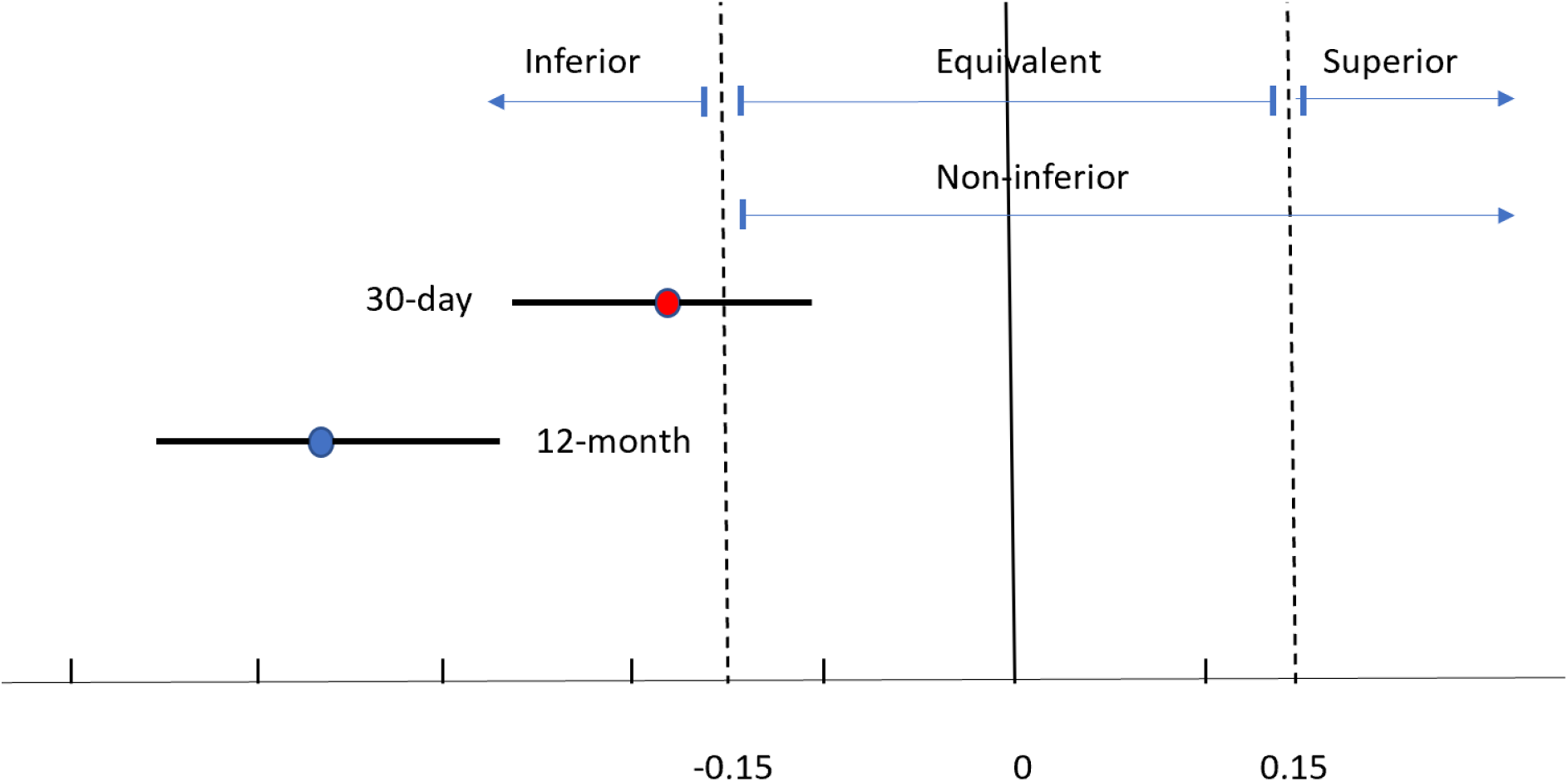
Treatment success proportional difference between operative management (OM) and non-operative management (NOM). Note. Non inferiority of NOM not demonstrated at 30 days or 12 months. Inferiority of NOM demonstrated at 12 months.

**Table 1:**
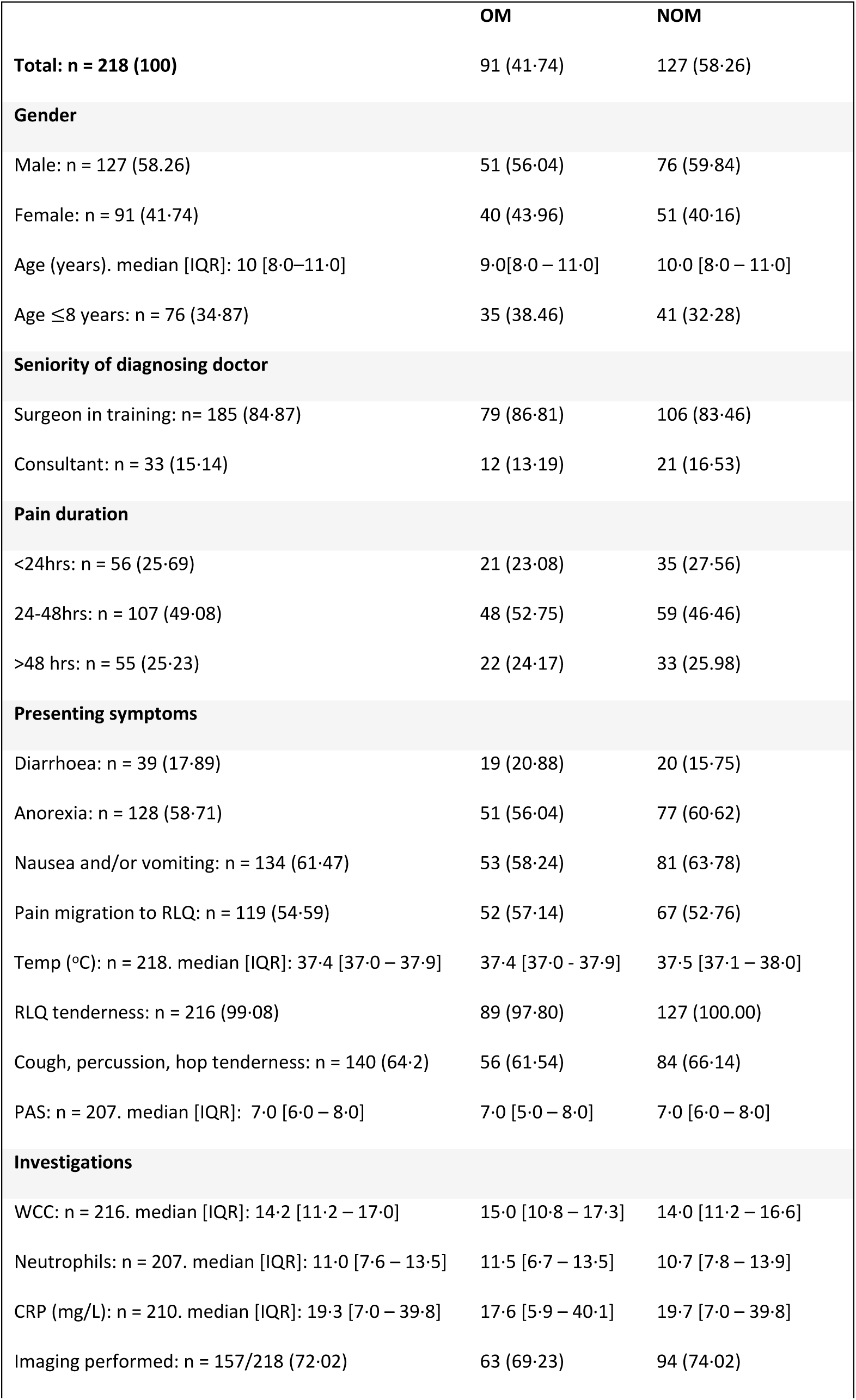
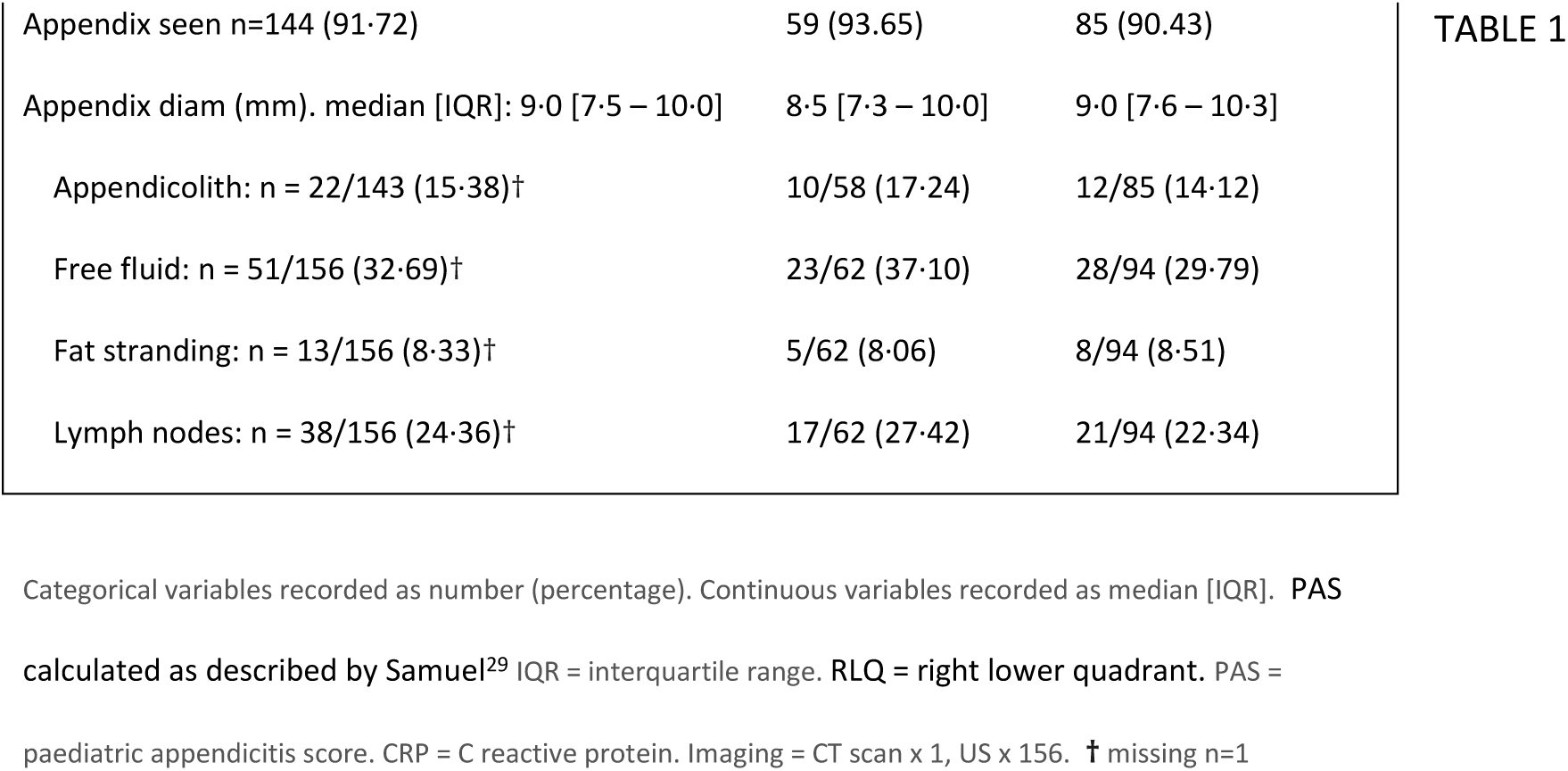
Comparison of children randomised to operative management (OM) vs non-operative management (NOM), excluding 30-day withdrawals (n=4).

|  | OM | NOM |
| --- | --- | --- |
| **Total: n = 218 (100)** | 91 (41.74) | 127 (58.26) |
| **Gender** |  |  |
| Male: n = 127 (58.26) | 51 (56.04) | 76 (59.84) |
| Female: n = 91 (41.74) | 40 (43.96) | 51 (40.16) |
| Age (years). median [IQR]: 10 [8.0–11.0] | 9.0[8.0 – 11.0] | 10.0 [8.0 – 11.0] |
| Age ≤8 years: n = 76 (34.87) | 35 (38.46) | 41 (32.28) |
| **Seniority of diagnosing doctor** |  |  |
| Surgeon in training: n = 185 (84.87) | 79 (86.81) | 106 (83.46) |
| Consultant: n = 33 (15.14) | 12 (13.19) | 21 (16.53) |
| **Pain duration** |  |  |
| <24hrs: n = 56 (25.69) | 21 (23.08) | 35 (27.56) |
| 24-48hrs: n = 107 (49.08) | 48 (52.75) | 59 (46.46) |
| >48 hrs: n = 55 (25.23) | 22 (24.17) | 33 (25.98) |
| **Presenting symptoms** |  |  |
| Diarrhoea: n = 39 (17.89) | 19 (20.88) | 20 (15.75) |
| Anorexia: n = 128 (58.71) | 51 (56.04) | 77 (60.62) |
| Nausea and/or vomiting: n = 134 (61.47) | 53 (58.24) | 81 (63.78) |
| Pain migration to RLQ: n = 119 (54.59) | 52 (57.14) | 67 (52.76) |
| Temp (°C): n = 218. median [IQR]: 37.4 [37.0 – 37.9] | 37.4 [37.0 - 37.9] | 37.5 [37.1 – 38.0] |
| RLQ tenderness: n = 216 (99.08) | 89 (97.80) | 127 (100.00) |
| Cough, percussion, hop tenderness: n = 140 (64.2) | 56 (61.54) | 84 (66.14) |
| PAS: n = 207. median [IQR]: 7.0 [6.0 – 8.0] | 7.0 [5.0 – 8.0] | 7.0 [6.0 – 8.0] |
| **Investigations** |  |  |
| WCC: n = 216. median [IQR]: 14.2 [11.2 – 17.0] | 15.0 [10.8 – 17.3] | 14.0 [11.2 – 16.6] |
| Neutrophils: n = 207. median [IQR]: 11.0 [7.6 – 13.5] | 11.5 [6.7 – 13.5] | 10.7 [7.8 – 13.9] |
| CRP (mg/L): n = 210. median [IQR]: 19.3 [7.0 – 39.8] | 17.6 [5.9 – 40.1] | 19.7 [7.0 – 39.8] |
| Imaging performed: n = 157/218 (72.02) | 63 (69.23) | 94 (74.02) |

|  |  |  |  |
| --- | --- | --- | --- |
| Appendix seen n=144 (91.72) | 59 (93.65) | 85 (90.43) | TABLE 1 |
| Appendix diam (mm). median [IQR]: 9.0 [7.5 – 10.0] | 8.5 [7.3 – 10.0] | 9.0 [7.6 – 10.3] |  |
| Appendicolith: n = 22/143 (15.38)† | 10/58 (17.24) | 12/85 (14.12) |  |
| Free fluid: n = 51/156 (32.69)† | 23/62 (37.10) | 28/94 (29.79) |  |
| Fat stranding: n = 13/156 (8.33)† | 5/62 (8.06) | 8/94 (8.51) |  |
| Lymph nodes: n = 38/156 (24.36)† | 17/62 (27.42) | 21/94 (22.34) |  |

Categorical variables recorded as number (percentage). Continuous variables recorded as median [IQR]. PAS
calculated as described by Samuel<sup>29</sup> IQR = interquartile range. RLQ = right lower quadrant. PAS =
paediatric appendicitis score. CRP = C reactive protein. Imaging = CT scan x 1, US x 156. † missing n=1

In the OM arm, CA was found in 10/90 (11.11%) and 5 (5.55%) had a normal appendix. One patient with SA had a 9 mm carcinoid tumour on histology. There were five complications in four patients (4.44%) (Table 2).

**Table 2:**
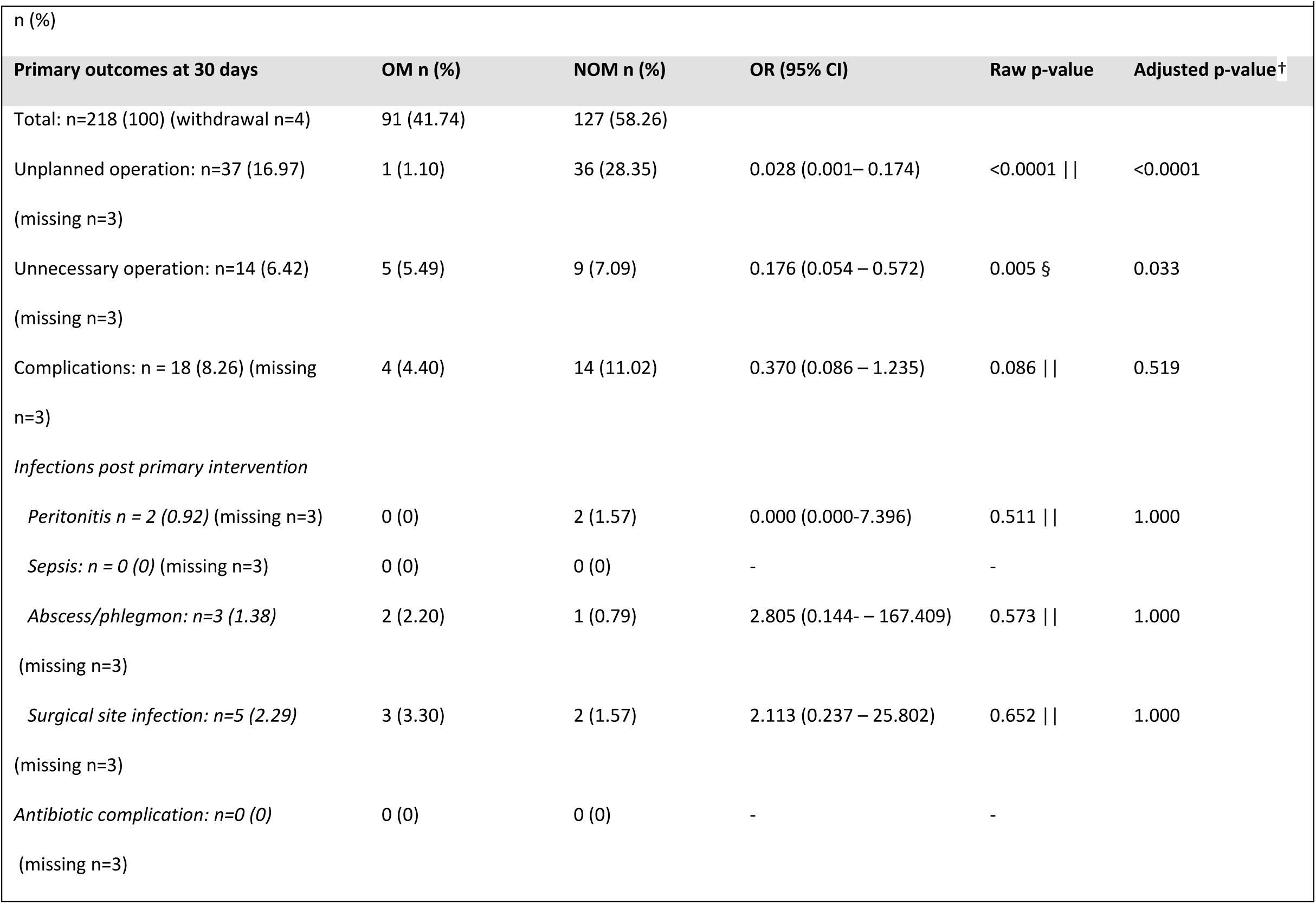
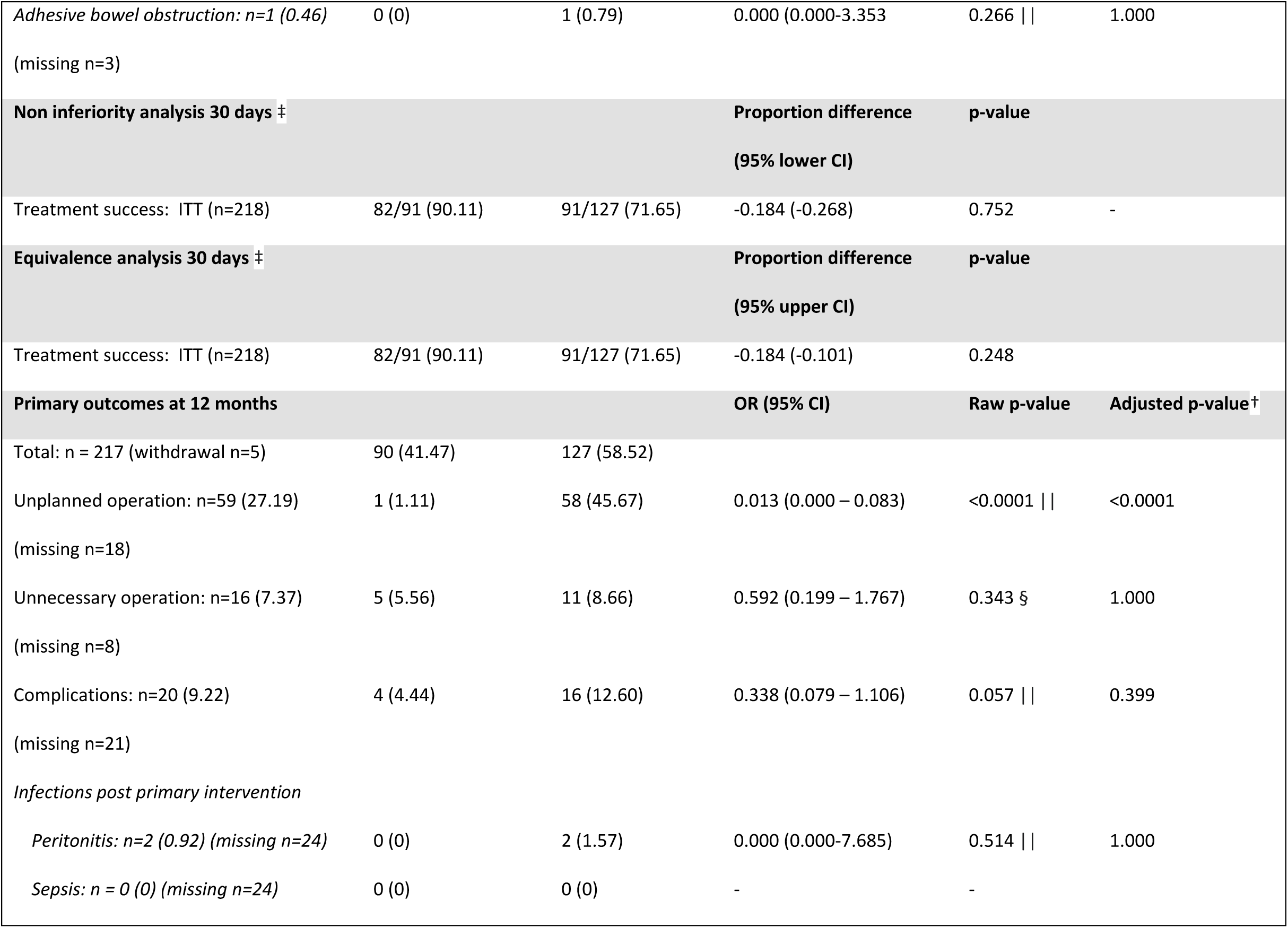
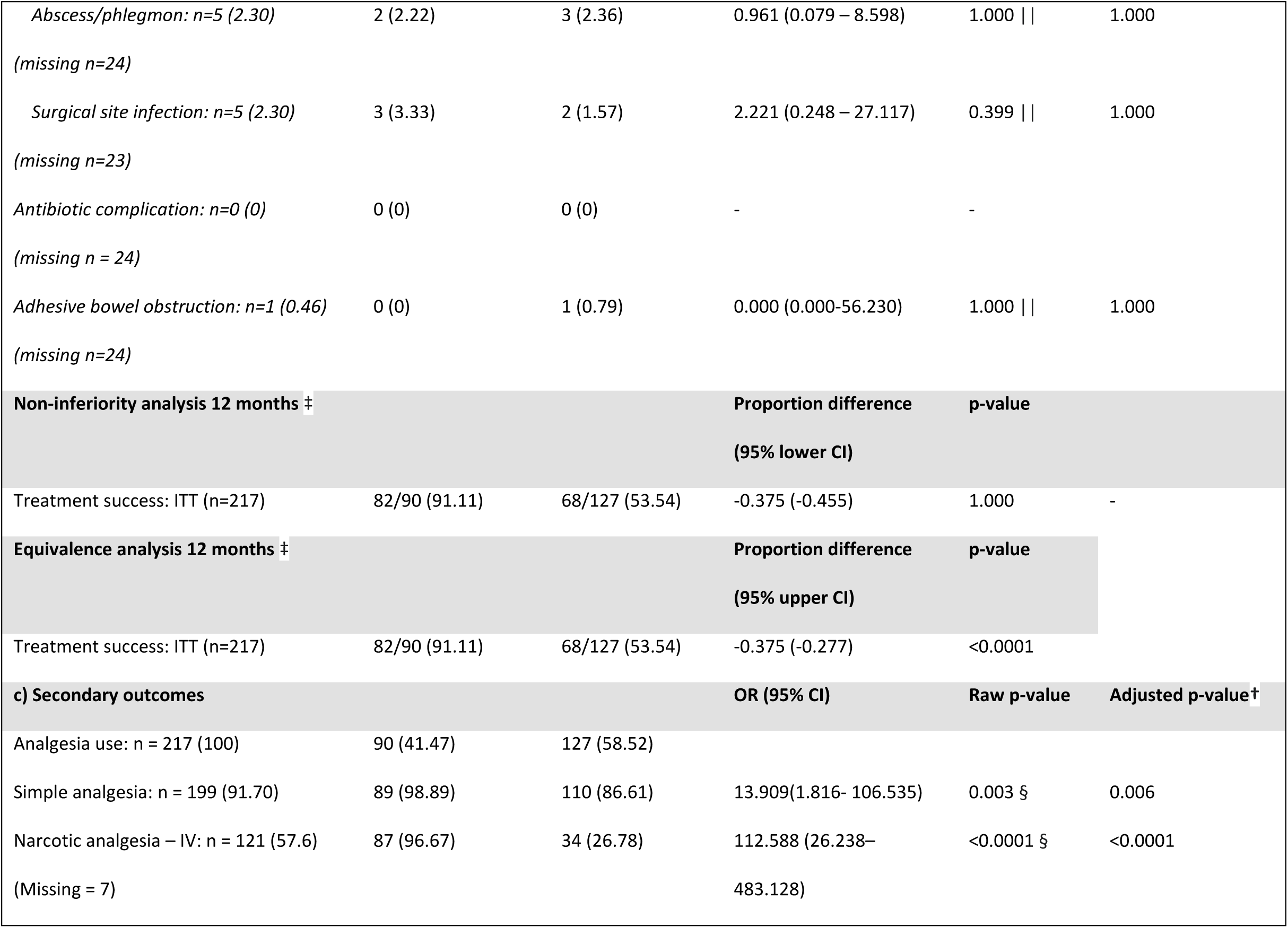
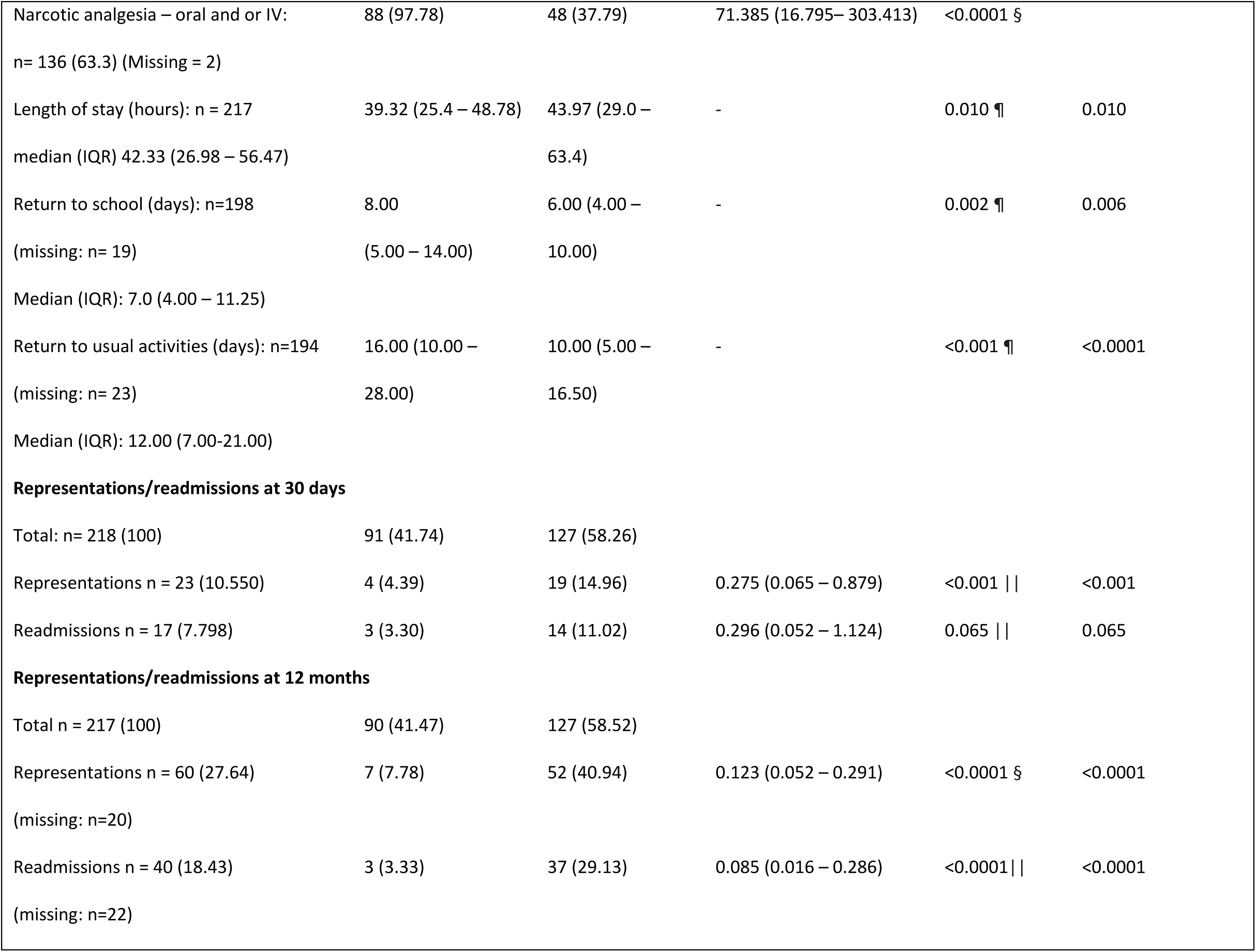
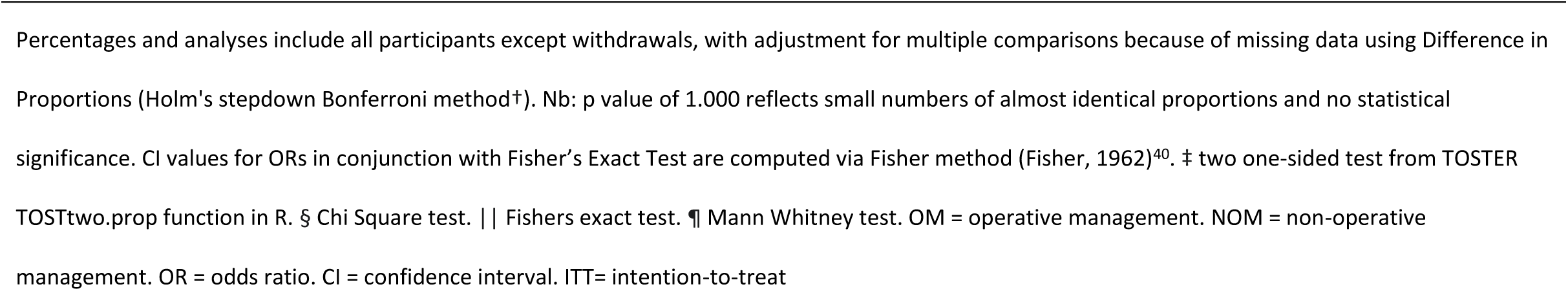
a) Primary outcomes at 30 days b) Primary outcome at 12 months c) Secondary outcomes.

In the NOM arm, 58/127 (45.67%) underwent an unplanned operation: 26 (44.83%) while still in hospital, 10 (17.24%) after discharge but within 30 days and 23 (39.66%) between 30 days and 12 months. Details are provided in Figure 4. There were no factors identified that correlated with requirement for an unplanned operation in the NOM group (Table S1).

**Figure 4:**
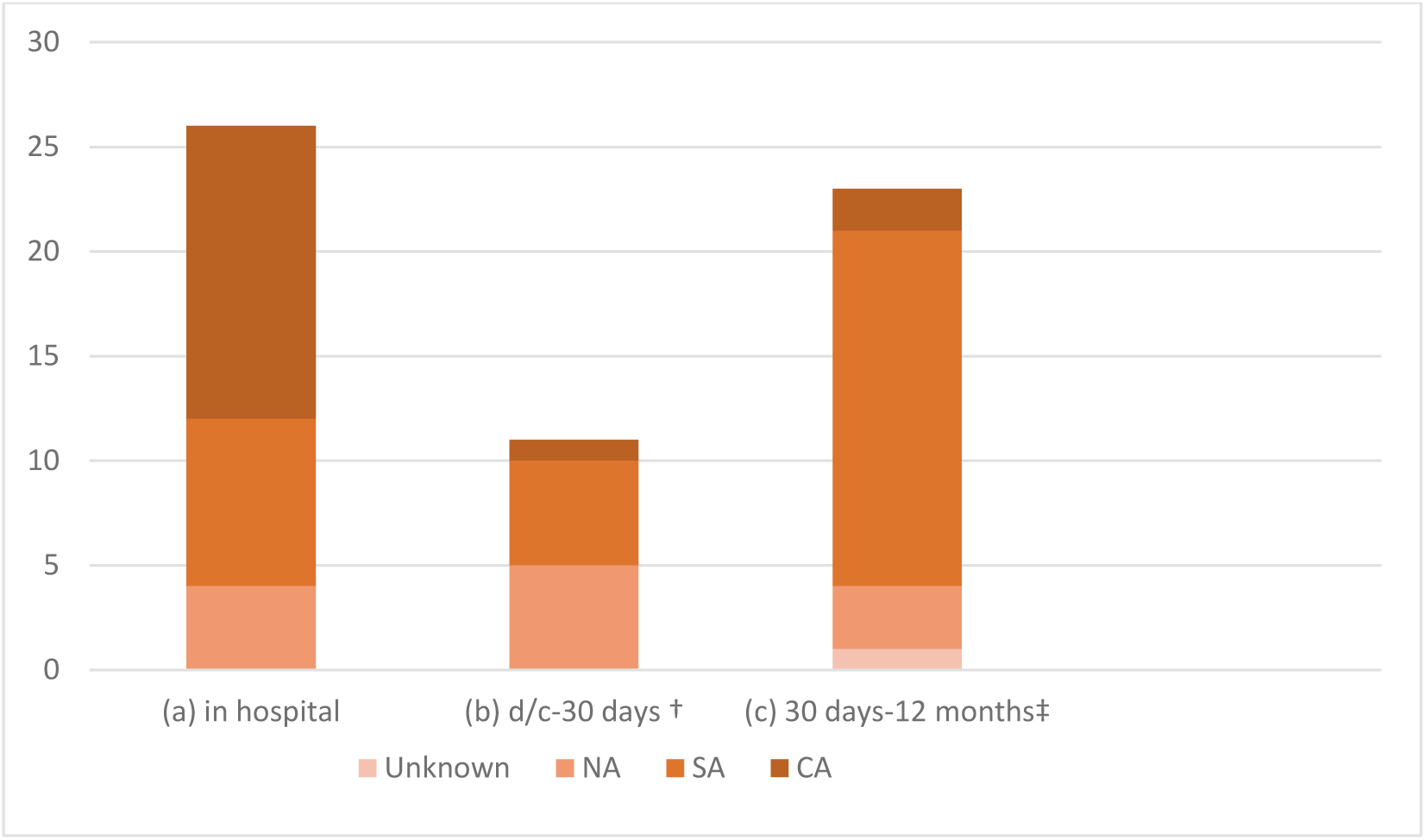
Unplanned operations in the non-operative arm. Note. NA= normal appendix. SA = simple appendicitis. CA = complicated appendicitis. Complicated appendicitis defined as appendicitis with perforation, fibro-purulent exudate or abscess formation and excluded gangrenous appendicitis.41 † The one patient who represented with complicated appendicitis at 3 days post discharge likely had a localised abscess at discharge. It was drained, followed by interval appendicectomy at 5 months. ‡One patient underwent appendicectomy at a different hospital for which findings were unavailable.

Secondary outcomes are summarised in Table 2. There was no significant difference in the number of complications at 30 days or 12 months. No child became septicaemic, there were no major antibiotic complications and no deaths. One child initially randomized to NOM but subsequently undergoing appendicectomy developed an adhesive bowel obstruction, which settled.

Narcotic analgesia was given significantly more frequently with OM and length of stay was significantly shorter. However, children randomized to NOM returned to school and usual activities significantly sooner.

By 12 months, 60 children (27.65%) had re-presented to the emergency department after discharge and 40 (18.43%) required readmission, significantly more in the non-operative arm (p<0.0001).

## Discussion

To our knowledge, this is the first adequately powered multicentre, randomized control trial comparing the outcomes of OM to NOM of appendicitis in children. Non inferiority was not demonstrated at either 30 days or 12 months: in fact, NOM was conclusively inferior at 12 months, with almost 50% of children initially randomised to NOM, subsequently undergoing appendicectomy. However, initial NOM as offered in this study, was safe, with no difference in complications between OM and NOM.

The study design was similar to Salminen’s 2015 landmark study in adults.^7^ While non-inferiority was not confirmed at 12 months, almost three quarters of those randomised to NOM were successfully treated without the need for operation, compared to only 50% in our study. This treatment failure rate may have been contributed to by the study design. Firstly, diagnostic imaging was not stipulated and secondly, the threshold for OM in the NOM arm was low.

Firstly, not requiring imaging meant cases of CA were inadvertently recruited. Most in the NOM arm were detected while still in hospital and likely were complicated at randomization. Adult studies have usually stipulated imaging confirmation, often with CT scan. Two of the small randomized controlled trials in children also required imaging confirmation – ultrasound or CT scan.^6,7^In our setting, cross-sectional imaging is not routine because of radiation concerns,^30^ and therefore was not part of the study design. Although ultrasound is safe, and has been shown to be useful in diagnosing CA in this age group,^31^ reliance on it can increase the incidence of negative appendicectomy in SA.^32^ There is a balance between the risk of over diagnosing SA and potentially using antibiotics (or operating) when no treatment is warranted, and the benefit of detecting CA before embarking on NOM.

Secondly, children in the NOM that may have settled given more time, underwent appendicectomy, with a 20% negative appendicectomy rate in this group (Figure 4), higher than the 10% base line at the trial sites. This may have been contributed to by study design, and surgeon, as well as carer factors. Nevertheless, a realistic threshold for offering appendicectomy after initial NOM, while contributing to failure to demonstrate non-inferiority, helps maintain safety and acceptability: equally important aspects of management to clinicians and families.

Looking at other paediatric literature, in Svensson’s 2015 pilot randomized controlled trial, treatment success at one year (with a similar definition to our study) was 15/24 (62%).^19^ Gorter’s 2018 interim report of 44 cases from a Dutch multicentre study^20^ excluded those with a faecolith, which has been associated with NOM treatment failure in adults^11^ and children,^34^ although not in ours (Table S1). Interestingly, Gorter^20^ who reported treatment success of 76%. looked at complications as the primary outcome. As in our study, they were equal for both OM and NOM. This prioritisation highlights the focus on safety when considering the place of NOM.

A recent, larger, non-randomized trial, allowing parents to choose treatment arm, also found a higher success for NOM at 12 months of 62%. As well as giving families agency in clinical decision making, this was likely also contributed to by tighter selection criteria.^35^

Longer mean length of stay was contributed to by the study design, as well as the longer stay of those undergoing subsequent appendicectomy. Prechal’s 2019 meta-analysis of adult studies showed no difference in length of stay,^14^ but studies in children vary on this measure, showing no difference^34^ or a shorter stay for OM.^33^ With the reassurance that initial NOM is safe, hospitalisation duration for NOM may be reduced.

Narcotic analgesia use was lower and return to school and usual activities earlier with NOM, both of the latter, by inference, corelated with the benefit of shorter interruption to work and other commitments of carers. Studies with similar follow-up periods concur, showing financial benefits^36^ and reduced disability days.^35^

At 12 months, re-presentation and readmission to hospital was higher for NOM. As well as treatment failure, the desire for such review may indicate a higher level of concern about the adequacy of treatment with antibiotics alone. This underlines the importance of involving families in decision making. Efforts to address this could include routine early follow-up calls and written discharge information.

Appendiceal carcinoid occurred in one patient in the operative arm, co-existent with SA. The reported incidence at appendicectomy is approximately 0·3%, with most not seen macroscopically.^37^While the slim possibility of an incidental carcinoid does not preclude the use of NOM, informed consent is required. Imaging to exclude a large carcinoid has also been recommended.^37^

Other issues to contemplate when considering NOM for SA include the lifetime risk of recurrence, and antibiotic stewardship. The lifetime risk of appendicitis is 7-8%.^1^ Longer follow-up over many years will be required to see if this risk is reduced or just delayed after NOM. The study design required a longer duration of antibiotics for NOM, shown to be associated with increased antibiotic complications in other settings.^38^ The risk of antibiotic resistance also needs careful thought.^39^ Choice to use NOM with antibiotics should only occur when OM would otherwise be pursued, resisting the temptation to speculatively treat borderline cases this way. Further study is needed to see if the duration of the antibiotic course for NOM could be safely reduced.

## Limitations

The study design meant some children were enrolled who had CA or a normal appendix. Requiring imaging may have improved treatment success.

Randomization was skewed towards NOM. This may be because sealed envelopes were not used in order, and repeat randomization was required for some stratification groups with higher enrolments, while randomizations in other groups were not exhausted. Nevertheless, recruiters were not aware of this skew. The higher numbers in intervention arm ensured the power of the study, while numbers in the control arm were in line with recruitment aims.

The requirement to change antibiotics during the trial was unavoidable if the study was to continue. Engaging the antibiotic stewardship teams to identify a suitable replacement helped to maintain consistency in antimicrobial profile.

Recruitment rate was just below the 30% projected in the original study protocol.^24^ and extended over more than three years. This was contributed to by the temporary trial suspension, low recruitment over holiday periods, availability of trial personnel and by parental preconceived ideas about what treatment they considered to be preferable. In addition, it is possible that discomfort with randomization to NOM, both for parents and surgeons, may have led to a higher rate of its treatment failure. The inability to blind an interventional study such as this means these confounders are difficult to avoid. A study design which allows parent choice, may boost participation and affect treatment outcomes, as in Minneci’s 2020 trial.^35^

## Conclusion

Non-inferiority of NOM for SA in children was not shown in this study. The challenges of comparing OM to NOM, even in an adequately powered trial were evident, including: selection of cases; biases introduced by the inability to blind; and the lack of equivalent outcome measures. In addition, the life-time risk of appendicitis after NOM has not been quantified – a more pivotal issue in children than adults.

However, NOM has been shown to be safe, associated with less narcotic analgesia use, quicker recovery and return to normal activities.

If NOM is to be offered outside of a study setting, consideration should be given to imaging confirmation, informed consent about published outcomes and the small risk of missed pathology such as carcinoid. Further research to refine the place of NOM for SA in children is required, including evaluation of longer-term outcomes and resource implications. Assessing the place of choice, and the acceptability of the treatment for children and their carers is important. The attitudes of surgeons and other health professionals are also likely to be of influence, and require evaluation.

## Data Availability

All data produced in the present work are contained in the manuscript and associated tables.

## Acknowledgements

The authors wish to acknowledge the contribution of the following people to the completion of this study.

UNSW Medical students who contributed to data collection

Kylie-ann Mallitt, Nancy Briggs and Andrew Kirk for their assistance with statistical methods and data analysis.

