## Supplemental Table 1 for "Non-operative management of uncomplicated appendicitis in children: a randomised, controlled, non-inferiority study evaluating safety and efficacy"

Table S1: A comparison of patients randomised to non-operative management who failed treatment by 12 months to those who did not.

| n (%) | Success | Failure | OR (95% CI) | p-value |
|---|---|---|---|---|
| Total: n = 127 (100) (missing n=7) | 68(53.54) | 59(46.46) | | |
| Gender | | | | |
| Female: | 27(39.71) | 25(42.37) | 0.896 (0.441-1.820) | 0.760 § |
| Male: | 41(60.29) | 34(57.63) | | |
| Age | | | | |
| 5-8 years: n = 34 (31.2) | 25(36.76) | 16(27.12) | 1.563 (0.733-3.330) | 0.246 § |
| 9-16 years: n = 75 (68.8) | 43(63.24) | 43(72.88) | | |
| Site | | | | |
| Site 1: n = 60 (55.0) | 38(55.89) | 35(59.32) | 0.869 (0.429-1.760) | 0.696 § |
| Site 2: n = 49 (45.0) | 30(44.11) | 24(40.68) | | |
| Diagnosing Doctor | | | | |
| Surgeon in training: n = 105 | 54(79.41) | 51(86.44) | 0.605 (0.234-1.563) | 0.297 § |
| Consultant: n = 22 | 14(20.59) | 8(13.56) | | |
| Presenting symptoms | | | | |
| Pain duration >48 hours: n = 33 | 18(26.47) | 15(25.42) | 0.947 (0.427-2.099) | 0.893 § |
| Diarrhoea: n = 20 | 11(16.18) | 9(15.25) | 1.072 (0.411-2.798) | 0.887 § |
| Anorexia: n = 76 | 38(55.89) | 38(64.41) | 0.700 (0.342-1.433) | 0.328 § |
| Nausea and/or vomiting: n = 81 | 40(58.82) | 41(69.49) | 1.594 (0.764-3.325) | 0.212 § |
| Pain migration to RLQ: n = 67 | 37(54.41) | 30(50.85) | 0.867 (0.431-1.743) | 0.688 § |
| Temperature ≥38 $^{0}$C: n = 33 | 15(22.06) | 18(30.51) | 1.551 (0.699-3.443) | 0.279 § |
| RLQ tenderness: n =127 (100) | 68(100) | 59(100) | - | - |
| cough/hop/perc tenderness: n = 84 | 44(64.71) | 40(67.80) | 1.148 (0.549-2.404) | 0.714 § |
| PAS: median (IQR) = 7.0 (6.0 – 8.0) n= 123 (missing n= 4 due to missing neutrophil count) | 7.0(6.0 – 8.0) | 7.0(6.0 – 8.0) | - | 0.556¶ |
| PAS > 6 (missing n=4 due to missing neutrophil count) | 35/64(54.69) | 37/59(62.71) | 1.394(0.677-2.867) | 0.367 § |
| Investigations | | | | |
| Leukocyte count >10 x $10^{9}$/l: n = 102 | 55(80.89) | 47 | 0.926 (0.386-2.223) | 0.863 § |
| Neutrophil count >7.5 x $10^{9}$/l: missing n=4 | 52/64(81.25) | 46/59(77.97) | 0.817 (0.339-1.967) | 0.651 § |
| CRP: median (IQR) = 18.70(7.0 - 39.775) n = 122 (missing n = 5) | 21.45(10.20-42.25) missing n=2 | 15.50(5.10-37.9) missing n=3 | - | 0.241¶ |
| Imaging on admission: n = 94 | 49(72.06) | 45 | 1.246 (0.560-2.774 | 0.589 § |
| Appendix Seen n= 85/94 | 45/49(91.84) | 40/45(88.89) | 0.711 (0.179-2.832 | 0.628 § |
| Appendiceal diam >6mm | 43/45(95.56) | 39/40(97.50) | 1.814 (0.158-20.796) | 0.628 § |
| Faecolith: n = 12/85 (missing n=9) | 6(12.24) | 6(13.33) | 1.147 (0.338-3.891) | 0.826 § |

For ITT analysis, the 7 patients for whom data is missing are assumed to have been treated successfully with NOM. ITT = intention to treat. RLQ = right lower quadrant. $^{0}$C = degrees Celsius. PAS = paediatric appendicitis score.
Imaging = 1 x CT scan and 87 x ultrasounds. § Chi Square test. ¶ Mann Whitney test
